# How early family activities predict life satisfaction among parents with adult children

**DOI:** 10.1101/2024.11.22.24317775

**Authors:** M. Nils Peterson, Kathleen Bordewieck, Elijah Velluti-Fry, Julia L. Jansson, Gwen E. Peterson, Tarla R. Peterson

**Affiliations:** Fisheries, Wildlife and Conservation Biology Program, Department of Forestry and Environmental Resources, North Carolina State University, Raleigh, NC, United States; Department of STEM Education, North Carolina State University, Raleigh, NC, United States; Department of Wildlife, Fish, and Environmental Studies, Swedish University of Agricultural Sciences, Umeå, Sweden

**Keywords:** aging, family, happiness, recreation, well-being

## Abstract

Life satisfaction represents a relatively universal social goal. Research suggests familiar interactions of diverse types may shape life satisfaction, but many questions remain about the valence and relative importance of variables shaping life satisfaction. In this exploratory study, we examined the relationships between frequency of shared parent-child activities in early life stages and self-reported parental life-satisfaction after children leave home using a case study of NC State University and University of Texas at El Paso students’ parents (n=92). Frequency of shared housework with young children was the most important, and positive, predictor of life satisfaction among parents after the children left home, and shared religious activities were also a positive predictor. Conversely, frequency of playing sports with young children was a negative predictor of later life satisfaction among parents. We did not detect a relationship for shared visits to local parks. Current activity level and income level both positively predicted life satisfaction, but we did not detect relationships for gender or marital status. This preliminary research highlights several novel ways shared family activities may affect later life satisfaction among older parents, but requires larger scale research to assess if an how findings apply in other contexts.

## Introduction

Global scale social and environmental challenges (e.g., climate change, disease, war) threaten human well-being worldwide and highlight the need for research factors that contribute to resilience and happiness (Myers and Patz, 2009; Beal et al., 2022). Well-being, however, is a complex concept shaped by social conditions (e.g., availability of education, food, housing, healthcare [Human Development Index]), self-realization (i.e., eudaimonic approaches), individual participation in practices associated with quality of life (e.g., exercise, healthy diets, education; Veenhoven, 1996), and people feeling well (Ruggeri et al., 2020). The concept is further complicated by intersubjectivity with experts assessing well-being for other people and individuals assessing their own well-being (i.e., subjective well-being; Sniezek et al., 2010). The latter division is often paired with different metrics of well-being where medical or economic data informs external assessments of well-being and survey data informs subjective well-being. Subjective well-being also includes temporal elements, with some measures (e.g., life satisfaction) ranging from an assessment of the current moment to a subject’s entire life (Veenhoven, 1996). Composite measures of well-being can be used to integrate some aspects of well-being, but 10 or more dimensions ranging from resilience to self-esteem may be needed to capture psychological well-being (Huppert and So, 2013; Rugeri et al., 2020).

Understanding the predictors of well-being appears worth the considerable effort required given well-being predicts a host of positive outcomes in addition to simply feeling well. For instance, well-being predicts increased workplace productivity, learning efficacy, creativity, collaboration, volunteerism, relationship quality, national economic performance, and physical health measures for individuals and nations (Deaton, 2008; Diener, 2012; Taris, 2018; Oishi, 2007). The drivers of well-being are complex, diverse, and differ based on context. Age exhibits non-linear relationships with life satisfaction, increasing until middle age, and then declining toward the end of life (Gerstorf et al., 2010). Well-being may be more resilient to the presence of illness among older people than younger people (Harrison et al., 2016). Income tends to relate positively with life satisfaction (Diener et. al., 2002), but both income inequality and national context can mediate the relationship (Harrison et al., 2016).

The relationship between employment and well-being illustrates the difficulty in determining causality, with students reporting the lowest well-being, retired people reporting the highest levels of well-being, and both employed and unemployed people reporting intermediate levels (Harrison et al., 2016). In addition to struggling with low well-being levels, students may be particularly vulnerable to declines in well-being after a disturbance (Jackson et al., 2021; Liverpool et al., 2023; Sheldon et al., 2021). Gender effects on well-being may have flipped between 1970 and 2000, with women shifting from higher to lower well-being than men during the period (Stevenson and Wolfers, 2009), and this trend may be exacerbated by large scale disturbances (e.g., COVID-19) that differentially affect women due to higher family responsibilities and loneliness (Etheridge and Spantig, 2022). Physical health and activity are among the strongest and most consistent predictors of well-being and life satisfaction (Fastame, 2021; Schmiedeberg and Schröder, 2017). All these effects may vary by nation and cultural context, with Nordic nations, especially Finland and Denmark, frequently ranking at the top of well-being and happiness scales (Helliwell et al., 2024; Harrison et. al., 2016). These national differences may in part reflect lower social inequality supporting both higher levels of subjective well-being and more resilient well-being.

Social and familial relationships clearly shape life satisfaction influencing both parents and children. Social network size and having more kin within social networks appear to drive higher life satisfaction if familial relationships are positive (Fuller-Iglesaias, et al., 2015). Shared family activities can shape well-being for both children and parents with shared meals and leisure typically increasing well-being and shared homework and housework decreasing well-being (Offer, 2013; Coyl-Sheperd and Hanlon, 2013; Offer, 2014). These relationships may persist into late life with overall life satisfaction positively related to satisfaction with family relationships, social leisure time, and religious well-being (Fastame, 2021; Schmiedeberg and Schröder, 2017). Contributions to life satisfaction from social interactions may reflect the human need for affiliation (La Guardia and Patrick, 2008).

We build on this research using a retrospective study to better understand how leisure and work activities that parents share with children may predict life satisfaction of parents later in life. We test the hypothesis that more frequent shared activities in four domains (household tasks, sports, religious activities, and visiting local parks) predict life satisfaction among parents of adult children, while controlling for income and current physical activity levels. We acknowledge that life satisfaction is clearly only one element of well-being, but an important one (Ruggeri et al., 2020). The concept hearkens back to the seminal question of how we can foster what humans consider a life well lived (Veenhoven, 1996; Diener et al., 2002). The four domains for shared activities we address are grounded in self-determination theory which posits humans have three fundamental psychological needs: autonomy (self-rule or sense of control), competence (being challenged in but also masterful in activities), and relatedness (needing interpersonal bonds; La Guardia and Patrick, 2008; Deci and Ryan, 2012). Each of the four shared activity domains may contribute to the need for relatedness by their nature of being shared and potentially building long term familiar bonds (Newman et al., 2014; Schmiedeberg and Schröder, 2017; Lim and Putnam, 2010). Household tasks, sports, and religious activities may also relate to competence and autonomy given they aim to achieve goals, and often build competence in some skills. Religious activities may influence life satisfaction by reducing uncertainty about identity and increasing personal sense of control (autonomy; Hayward and Elliott, 2014).

## Methods

A convenience sample was used to survey parents who had children in college. Parent emails were requested from students in a Human Dimensions of Wildlife Management class at North Carolina State University (n = 78 students) and a Methods of Research in Communication Studies class at The University of Texas at El Paso (n = 38). Students randomly selected one of their parents to invite to participate. A parent of all 78 North Carolina State University students participated, and a parent of 14 students at the University of Texas at El Paso participated (31.58% participation rate). Data were collected via an online survey on the Qualtrics platform (Qualtrics, Provo, UT). Each individual was given a unique survey link, which was open between March 28^th^ and April 18^th^, 2018. The initial email was sent on March 28^th^, 2018, and participants received a follow-up reminder email on April 4^th^ and April 11^th^ of 2018.

Life satisfaction was measured using Diener et al.’s (1985) five-item Satisfaction with Life Scale with each item ranging from “strongly disagree” (1) to “strongly agree” (7) (Table 1). This yielded a scale with a minimum of 5 and a maximum of 35. The frequency of parent-child activities (play sports outside with my children, work on household tasks [inside and outside] with my children, participate in religious activities with my children, visit local parks with my children) was measured on a five-point scale ranging from “never” (1) to “once a week or more” (5). All respondents were asked to indicate their age, marital status (converted to a dummy variable for married or not), gender (female, male, or other), current state of residence, age range of their children, highest education level achieved (less than high school, high school/GED, vocational or trade school, associate’s degree, bachelor’s degree, graduate or professional degree), self-identified ethnicity, and annual household income (Less than $25,000; $25,000 - $50,000; $50,001-$75,000; $75,001-100,000; $100,001-$125,000; $125,001-$150,000; Greater than $150,001), and current activity level (how many days from the last 7 in which they walked for at least 10 minutes at a time). The instrument was pretested by undergraduate volunteers (n = 67) at North Carolina State University. We also conducted cognitive interviews with a convenience sample of non-student adults and discussed each question to understand how interviewees perceived and interpreted questions and to identify and correct any wording or comprehension issues (n = 9; Drennan, 2003).

**Table 1.** Exploratory factor analysis for Satisfaction with Life scale items and overall Satisfaction with Life scale score among participants (*N* = 92)

| Item | Mean (SD) | factor loadings |
| --- | --- | --- |
| **Satisfaction with Life scale score** | **24.83 (5.98)** |  |
| In most ways my life is close to my ideal | 4.92 (1.51) | 0.87 |
| The conditions of my life are excellent | 4.98 (1.55) | 0.84 |
| I am satisfied with my life | 5.43 (1.42) | 0.91 |
| So far, I have gotten the important things I want in life | 5.55 (1.33) | 0.57 |
| If I could live my life over, I would change almost nothing | 4.16 (1.66) | 0.47 |
| Eigenvalue |  | 3.19 |
| % of variance explained |  | 57.06 |
| Cronbach's alpha |  | 0.85 |

We modeled how frequency of participation in each of the family activities predicted life satisfaction among parents with adult children using ordinary least squares regression. The model predicted the Satisfaction with Life Scale (Diener et al., 1985) using 9 independent variables (Table 2). We capped the model at 9 predictors given the evidence that *n* = 8-10 is required per variable for a robust regression model (Gotelli and Ellison, 2004; Jenkins and Quintana-Ascencio, 2020). We calculated variance inflation factors (VIFs), and scores were ≤ 1.53 for all variables suggesting multicollinearity problems were unlikely. We calculated the least significant number (LSN) of observations required to create significant test results at alpha 0.05 for each variable to highlight where future research with larger sample sizes may find effects not detected in this study. We report partial Eta Squared (η_p_ ^2^) values for each independent variable to assess effect size and adopt the Miles and Shevlin (2001) rules of thumb for small (η_p_ ^2^ ≥ 0.02), medium (η_p_ ^2^ ≥ 0.13), and large (η_p_ ^2^ ≥ 0.26) effect sizes in regression. All analyses were conducted using JMP Pro 17. The research was conducted according to the principles in the Declaration of Helsinki, and approved and ruled exempt by the North Carolina State University Institutional Review Board (Protocol Number 12531). Informed consent was obtained by participants checking a box on the survey indicating they had read the informed consent content and agreed to participate. Signatures were not collected because they would have created the only link to participants in otherwise anonymous data.

**Table 2.**
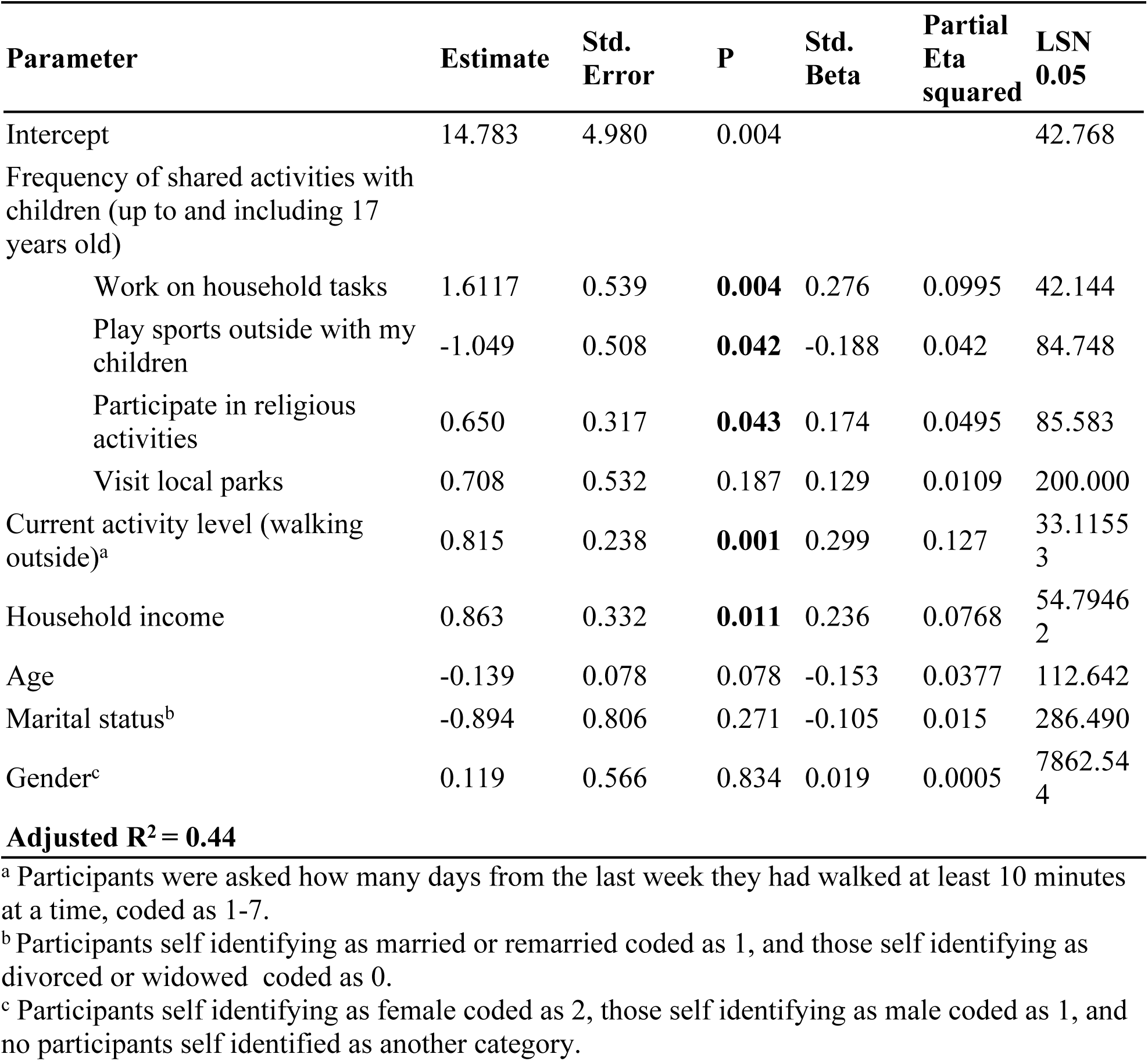
Ordinary least squares regression results estimating satisfaction with life among parents of college students based on frequency of shared activities with those children when they were still living at home, current activity levels among parents, and demographic variables of parents (N = 91).

## Results

A total of 92 parents of college-age children participated in the study. Of those, 56 resided in North Carolina, 8 chose not to share where they lived, 5 each resided in California and Texas, 4 reported currently living outside of the United States, 3 resided in Massachusetts, 2 each resided in New Jersey, Ohio, and Virginia, and 1 each resided in Maryland, Minnesota, New York, South Carolina, and Nevada. Most respondents reported being married (86%), and most reported being female (female = 64%, male 36%). Average age was 54.23 (SD = 6.59). Average household income before taxes (4.82, SD = 1.63) corresponded to slightly over $100,000. Average level of formal education was 4.87 (SD = 1.26), corresponding to a bachelor’s degree. Participants reported having on average 2.52 (SD = 1.10) children, with the oldest child of respondents averaging 25.70 (SD = 6.94) years old and the youngest child averaging 20.45 (SD = 7.53) years old. Most participants self-identified as White (83%) followed by Hispanic (11%), and Black (2%).

Exploratory factor analysis suggests the five Satisfaction with Life Scale items grouped onto one factor (Eigenvalue = 11.638 [all remaining values < = 0.258]) explaining 57.1% of variance and displaying high internal reliability and acceptable factor loadings for each item (Table 1). Mean satisfaction with life (24.83) corresponded with one unit above neutral or choosing ‘slightly agree’ for items in the scale (Table 1). The model (Table 2) had acceptable predictive capacity (adjusted R² = 0.45, n = 91). How often parents shared household tasks with their children during childhood (η_p_ ^2^ = 0.10) and parents’ current physical activity levels (η_p_ ^2^ = 0.13) were the strongest predictors of participants’ life satisfaction. Both were positive predictors with moderately large effect size (Table 2). The frequency of playing sports outside with children predicted lower life satisfaction, but the effect size was almost half that of completing household tasks, and relatively low (η_p_ ^2^ = 0.04; Table 2). Shared religious activities with children during their childhood years was a positive predictor of life satisfaction, but also less important than shared household tasks and had a small effect size (η_p_ ^2^ = 0.05). We did not detect effects for frequency of visiting local parks with children, and the LSN 0.05 suggests a relatively small sample size of 200 would have allowed detecting a positive effect (Table 2). Household income was positively associated with life satisfaction (Table 2). Age and unmarried status had negative coefficients with effect sizes detectable with reasonably small sample sizes (n < 300), but there was little evidence for a gender effect (Table 2).

## Discussion

Several factors may help explain why sharing household tasks with children was the strongest predictor of life satisfaction among parents after those children had gone to college. Household tasks occur regularly, perhaps daily, and thus may be more conducive to building long term familial bonds, and thus relatedness, than shared activities that are less common, as would be the case for nearly all leisure activities (Schmiedeberg and Schröder, 2017). Similarly, shared household tasks may render children more competent in household work, and provide parents, particularly mothers, more autonomy (Tomanovic, 2003). Further, shouldering too much household work as an individual renders outside employment less beneficial to having a sense of control (Ross and Mirowsky, 1992), so sharing household tasks with children may help parents feel more in control than they would otherwise. We are aware that most research in this domain suggests well-being and life satisfaction suffer from too much household work and imbalances in division of household tasks (e.g., González, et al., 2023; Ahn and Yoo, 2022; Carlson et al., 2020; Owoo and Lambon-Queyefio, 2021; Peristera et al., 2018). We, however, note that this relationship is nuanced. Housework can contribute to happiness through goal satisfaction and activity enjoyment, for example, when tasks align with key personal characteristics and activity preferences (Lee and Tang, 2022). Some research suggests unpaid housework may negatively impact women’s mental health with limited effects for men (Leopold, 2019; Ervin et al., 2022). It seems reasonable that enjoyable and shared household tasks may have positive impacts on long term life satisfaction for parents. Given satisfaction with housework can be a primary contributor to overall life satisfaction (Pagan, 2020), our results suggest that additional research exploring *how* shared housework among parents and children may contribute to their life satisfaction is warranted.

Given religious activity has long been among the most important predictors of life satisfaction in nations where religion is widely practiced (Domínguez and López-Noval, 2021; Hayward and Elliott, 2014; Sholihin et al., 2022), it is perhaps not surprising that frequency of shared religious activities between parents and children also predicted parental life satisfaction after children leave the home. This intuitive yet novel finding may also relate to self-determination theory. Specifically, religious activities may influence life satisfaction by reducing uncertainty about identity and increasing personal sense of control (i.e., autonomy; Hayward and Elliott, 2014), and these dynamics may have more influence on long term life satisfaction among parents when shared with adult children. The tendency for individual religious observance to promote health and well-being enhancing behaviors, including reduced cigarette and alcohol consumption and increased compliance with medical directives (Deaton and Stone, 2013; Hayward and Elliott, 2014), may also occur when religious observance is shared between parents and children. Future research with larger and more diverse samples would be needed to disentangle the contributions of individual observance and household level observance.

We were surprised by the frequency of shared sports activities with children predicting lower life satisfaction among parents after their children went to college, given generally positive associations between leisure and various forms of subjective well-being. Newman et al. (2014) suggested leisure research indicated these positive associations were mediated by affiliation (e.g., building social ties), autonomy (e.g., maintaining a leisure activity as one ages), detachment (e.g., escaping from pressure filled activities), meaning (e.g., providing purpose in life), and mastery (e.g., experiencing high skill). Playing sports with children may fail to generate life satisfaction through these mechanisms for multiple reasons. First, it may decrease the time available to create emotionally rewarding relationships outside the family (e.g., leisure well-being model; Carruthers and Hood, 2007) for parents where participation in other leisure activities including league sports might increase such social ties (Kim et al., 2020). Similarly, playing sports with children often involves sports of interest to the child, and can thus run counter to life enjoyment emerging from spontaneously following an inner interest for the parents (Newman et al., 2014). Finally, playing sports with children seems an unlikely path to developing high skill or mastery in a sport for most parents.

Small sample size may explain our failure to detect a positive relationship between visiting local parks with children and life satisfaction among parents after children went to college (i.e., n = 200 would have detected such a relationship). The relationship between time and nature and various forms of well-being is also complex, and questions remain about the duration of effects, dosage levels required to create an effect, and differential effects from different types of parks and activities within them (Beall et al., 2022; Peterson et al., 2024). The demographic control variables exhibited patterns lending face validity to the study since they largely reflect the extant literature. Specifically, current activity levels and income levels tend to be positive predictors of well-being and life satisfaction (Deaton, 2008; Deiner et al. 2002; Fastame, 2021), whereas age and marital status have less clear and mixed effects (Bartram, 2021; Deiner et al., 2002).

Future research is needed to address several limitations of this study. Larger studies would be able to detect more socially meaningful relationships and assess the potential for cultural, geographic, and economic variables to moderate relationships between parent-child activities and later life satisfaction among parents. Panel studies are needed to avoid recall bias associated with the retrospective pretest methods used in this study. Finally, research with more diverse populations is needed to understand how shared household tasks shape life satisfaction in contexts where those tasks are more likely to encroach on leisure and education. Despite these limitations, this study contributes to life satisfaction literature by documenting the intriguing possibility that several shared family activities, including shared household work, can shape long term life satisfaction.

## Data Availability

All relevant data are within the manuscript and its Supporting Information files.

## Acknowledgements

We would like to thank the families who participated in this study for volunteering their time. This research did not receive any specific grant from funding agencies in the public, commercial, or not-for-profit sectors.

